## Supplementary Tables & Figures for "Short-term acute exposure to wildfire smoke and lung function in a Royal Canadian Mounted Police (RCMP) cohort"

**List of supplementary Tables and Figures**

1. **Supplementary Table S1:** Component (unrotated) loading matrix of the principal component analysis (PCA)
2. **Supplementary Figure S1:** Scree plot of the principal component analysis
3. **Supplementary Figure S2:** Component loading plot (Bi-plot) of the PCA
4. **Supplementary Table S2:** Changes in lung function in association with per interquartile (IQR) change in air pollution index
5. **Supplementary Table S3:** Effect modification by smoking, any airway obstruction, childhood smoke exposure, and any parental lung disease on the association between the air pollution index (per interquartile change) and lung function indices of the RCMP officers.

**Supplementary Table S1:** Component (unrotated) loading matrix of the principal component analysis (PCA)

| **Variables** | **Comp 1** | **Comp 2** | **Comp 3** | **Comp 4** | **Comp 5** | **Comp 6** | **Comp 7** |
| --- | --- | --- | --- | --- | --- | --- | --- |
| **CO** | 0.39 | -0.14 | -0.41 | -0.15 | 0.04 | 0.09 | -0.79 |
| **CH_4_** | 0.35 | 0.62 | 0.32 | 0.44 | 0.15 | 0.41 | 0.04 |
| **NO** | 0.38 | 0.38 | -0.06 | -0.75 | -0.38 | 0.08 | 0.01 |
| **NO_2_** | 0.40 | -0.14 | 0.05 | 0.43 | -0.66 | -0.40 | 0.20 |
| **O_3_** | 0.39 | -0.05 | 0.47 | -0.18 | 0.49 | -0.59 | -0.11 |
| **SO_2_** | 0.35 | -0.65 | 0.33 | -0.12 | 0.01 | 0.55 | 0.16 |
| **PM_2.5_** | 0.38 | -0.02 | -0.64 | 0.04 | 0.40 | -0.04 | 0.54 |

Values are in μg/m^3^.

Abbreviations: CO: carbon monoxide; CH_4_: methane; NO: nitric oxide; NO_2_: nitrogen dioxide; O_3_: ozone; SO_2_: sulfur dioxide; PM_2.5_: particulate matter of aerodynamic diameter ≤2.5μm.

**Supplementary Figure S1:** Scree plot of the principal component analysis

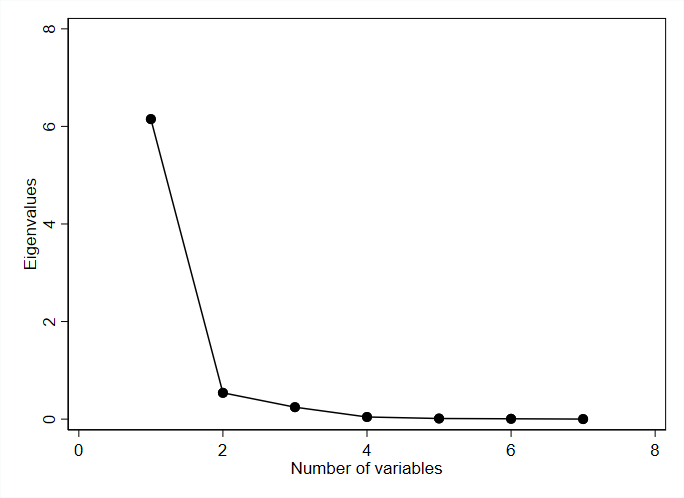

**Supplementary Figure S2:** Unrotated component loading plot (Bi-plot) of the PCA

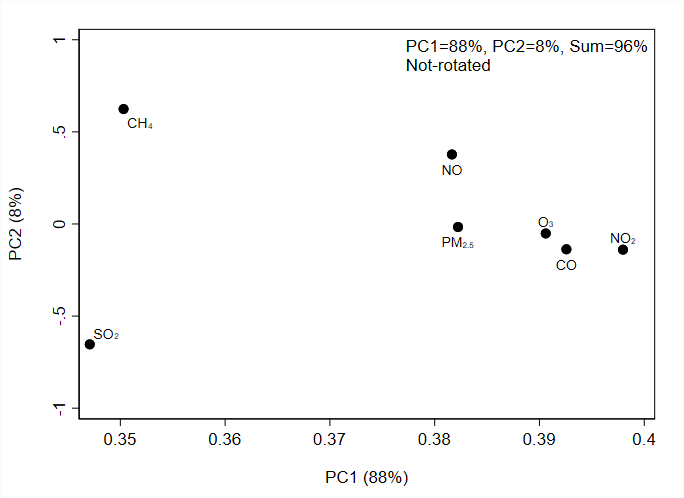

Abbreviations: CO: carbon monoxide; CH_4_: methane; NO: nitric oxide; NO_2_: nitrogen dioxide; O_3_: ozone; SO_2_: sulfur dioxide; PM_2.5_: particulate matter of diameter ≤2.5μm.

**Supplementary Table S2:** Changes in lung function in association with per interquartile (IQR) change in air pollution index

| **Variables** | **β (95% CI)** | | |
| --- | --- | --- | --- |
|  | **Model-1** | **Model-2** | **Model-3** |
| FVC (% predicted) | 0.36 (-0.80, 1.52) | 0.29 (-0.85, 1.43) | 0.30 (-0.84, 1.45) |
| FEV_1_ (% predicted) | 0.37 (-0.79, 1.53) | 0.35 (-0.77, 1.48) | 0.36 (-0.77, 1.48) |
| FEV_1_/FVC (%) | -0.20 (-0.75, 0.35) | 0.01 (-0.50, 0.52) | 0.005 (-0.51, 0.52) |
| TLC (% predicted) | 0.50 (-0.54, 1.54) | 0.43 (-0.60, 1.45) | 0.43 (-0.60, 1.46) |
| RV (% predicted) | 1.76 (-0.06, 3.57) | 1.55 (-0.28, 3.37) | 1.56 (-0.26, 3.37) |
| RV/TLC (%) | 0.40 (-0.05, 0.85) | 0.28 (-0.12, 0.67) | 0.28 (-0.12, 0.67) |

Data presented as linear regression coefficient (β) and 95% confidence interval, unless otherwise specified. Coefficients were calculated in respect to per IQR change (μg/m^3^) of the principal component (PC). Model-1: unadjusted model; Model-2: adjusted by age, sex, BMI, race, and smoking history. Model-3: Model-2 + PPE use (yes/no).

Abbreviations: BMI: body mass index; FEV_1_: forced expiratory volume in 1 second; FVC: forced vital capacity; TLC: total lung capacity; RV: residual volume; PPE: personal protective equipment.

**Supplementary Table S3:** Effect modification by smoking, any airway obstruction, childhood smoke exposure, and any parental lung disease on the association between the air pollution index (per interquartile change) and lung function indices of the RCMP officers

| **Effect modifier** |  | **FVC (% predicted)** | **FEV_1_ (% predicted)** | **FEV_1_/FVC (%)** | **TLC (% predicted)** | **RV (% predicted)** | **RV/TLC (%)** |
| --- | --- | --- | --- | --- | --- | --- | --- |
| Smoking* | No (N=174) | 0.55 (-0.75, 1.85) | 0.35 (-0.93, 1.63) | -0.13 (-0.71, 0.45) | 0.65 (-0.52, 1.82) | 1.82 (-0.18, 3.82) | 0.30 (-0.12, 0.71) |
|  | Yes (N=42) | -0.35 (-3.10, 2.39) | 1.11 (-1.61, 3.84) | 0.76 (-0.40, 1.93) | 0.13 (-2.35, 2.60) | 2.25 (-3.04, 7.53) | 0.63 (-0.60, 1.87) |
|  | *P^#^* | 0.32 | 0.95 | 0.30 | 0.35 | 0.56 | 0.89 |
| Any airway obstruction | No (N=149) | 0.03 (-1.27, 1.32) | 0.03 (-1.20, 1.25) | -0.13 (-0.60, 0.35) | 0.34 (-0.78, 1.47) | 1.62 (-0.36, 3.60) | 0.30 (-0.14, 0.73) |
|  | Yes (N=66) | 1.74 (-1.21, 4.68) | 1.61 (-1.37, 4.60) | 0.09 (-1.39, 1.57) | 1.42 (-1.31, 4.15) | 3.23 (-1.43, 7.90) | 0.53 (-0.44, 1.50) |
|  | *P^#^* | 0.22 | 0.24 | 0.73 | 0.35 | 0.48 | 0.70 |
| Childhood smoke exposure | No (N=87) | 0.93 (-0.78, 2.64) | 0.36 (-1.19, 1.91) | -0.46 (-1.12, 0.20) | 0.94 (-0.59, 2.47) | **2.71 (0.11, 5.30)** | 0.48 (-0.04, 0.99) |
|  | Yes (N=131) | -0.37 (-2.00, 1.24) | 0.35 (-1.32, 2.03) | 0.50 (-0.27, 1.26) | -0.19 (-1.64, 1.27) | 0.16 (-2.49, 2.82) | 0.05 (-0.55, 0.65) |
|  | *P^#^* | 0.26 | 0.91 | 0.09 | 0.26 | 0.18 | 0.34 |
| Parental lung disease | No (N=185) | 0.30 (-1.11, 1.71) | 0.43 (-0.90, 1.76) | 0.01 (-0.60, 0.63) | 0.60 (-0.68, 1.88) | 2.10 (-0.17, 4.38) | 0.37 (-0.12, 0.85) |
|  | Yes (N=33) | 0.56 (-1.39, 2.51) | 0.57 (-1.80, 2.95) | 0.11 (-0.93, 1.16) | 0.16 (-1.44, 1.75) | -0.03 (-0.293, 2.88) | -0.06 (-0.78, 0.65) |
|  | *P^#^* | 0.82 | 0.89 | 0.83 | 0.78 | 0.38 | 0.42 |

Data presented as linear regression coefficient (β) and 95% confidence interval, unless otherwise specified. Coefficients were calculated in respect to per IQR change (μg/m^3^) of the principal component (PC) adjusting for age, gender, BMI, race, and smoking history (*except for Smoking). P^#^: p value for interaction. Abbreviations: BMI: body mass index; FEV_1_: forced expiratory volume in 1 second; FVC: forced vital capacity; TLC: total lung capacity; RV: residual volume.
